# Interstitial Lung Diseases in a Longitudinal National Cohort of Veterans with Connective Tissue Diseases

**DOI:** 10.1101/2025.09.19.25336031

**Authors:** Mavi Rivera, Madiha Ahmad, Ayesha Iqbal, Mohleen Kang

**Affiliations:** Division of Rheumatology, Emory University School of Medicine, Atlanta, GA; Atlanta VA Health Care System, Decatur, GA; Division of Pulmonary, Allergy, Critical Care, and Sleep Medicine, Emory University School of Medicine, Atlanta, GA

## Abstract

**Introduction:** Interstitial lung diseases (ILDs) are a common manifestation in certain connective tissue diseases (CTD). Our study aims to describe the prevalence and time to diagnosis of CTD-ILD in veterans.

**Methods:** We used the Veterans Affairs Corporate Data Warehouse to select veterans with a) age ≥18 years, b) ≥ two ICD9 or ICD10 codes for Rheumatoid Arthritis (RA), Systemic Lupus Erythematosus, Systemic Sclerosis (SSc), Sjogren’s, Dermatomyositis/Polymyositis, Overlap, and Other CTD separated by ≥6 months from 1999 to 2022, c) rheumatology visit within 6 months. ILD was identified by ≥ one ICD9 or ICD10 code for ILD one year before CTD diagnosis or any time after. Kaplan-Meier curves and Cox regression analyses were performed.

**Results:** 88,559 veterans with CTD were identified with 8.8 (+/− 5.9) years of follow up. The majority were White males with RA and an average age of 60.4 years. 8,498 patients (9.6%) were diagnosed with ILD. 29% of veterans with CTD developed ILD within 1 year. Average time to diagnosis of ILD was 5.38 (SD 5.61) years. 42% of veterans with SSc were diagnosed with ILD while only 8% of veterans with RA developed ILD. The hazard of diagnosis of ILD in veterans with RA was 0.14 times compared to SSc (95%CI 0.13-0.15) adjusted for age, race, gender, smoking status, and CTD type.

**Conclusions:** In veterans, RA-ILD was the most common subtype, but veterans with RA were less likely to develop ILD compared to SSc. These results can influence strategies for ILD screening in patients with CTD.

**Significance and Innovations:**

- The prevalence of ILD in a national cohort of veterans with CTD was 9.6%.
- The prevalence of ILD in veterans with CTD was highest in SSc (42%) and lowest in RA at 8%.
- 29% of veterans with CTD developed ILD within +/− 1 year timeframe of CTD diagnosis.
- The average time to diagnosis of ILD after a diagnosis of CTD is approximately 6 years.

## Introduction

Connective tissue diseases (CTD) are a heterogeneous group of autoimmune disorders characterized by multisystem inflammation.^1^ Pulmonary involvement is common, specifically interstitial lung disease (ILD), and is a major cause of morbidity and mortality.^1,2^ The risk of development of ILD varies, even within a particular CTD, making it difficult to provide high quality evidence based guidelines for screening and early detection.^3–6^ ILD prevalence has been reported as highest in patients with Systemic Sclerosis (SSc) and lowest in Systemic Lupus Erythematosus (SLE).^7^ Moreover, the time to the onset of ILD is known to vary among the different CTDs, which may partly be influenced by differences in diagnostic practices and clinical surveillance.^1,7^ With development of high-resolution computer tomography (HRCT) scans, it has become possible to detect ILD early, but there is limited evidence to guide the timing and frequency of screening for ILD in patients with CTD. ^4–6,8,9^

Recent guidelines from the American College of Rheumatology (ACR)/American College of Chest Physicians (CHEST), American Thoracic Society (ATS) and European Respiratory Society (ERS)/European Alliance of Associations for Rheumatology (EULAR) recommend screening and monitoring for ILD in patients with CTD.^4–6^ These are largely conditional recommendations for screening high-risk patients with CTDs at disease presentation based on low certainty of evidence.^4–6^ The guidelines also acknowledge the lack of data to guide the frequency of rescreening and does not specify when the screening should stop if patients are stable.^4–6^ The US veterans present a unique population at risk for developing CTD and ILD due to their military exposures such as burn pit and presence of comorbidities such as PTSD.^10–13^ However, the prevalence of CTD and CTD-ILD in veterans is not well described except for RA-ILD.^14,15^ The goal of this study is to describe the prevalence of CTD and CTD-ILD and the time to diagnosis of CTD-ILD in a nationwide cohort of veterans.

## Methods

Emory University School of Medicine and the Atlanta VA Institutional Review Boards approved this study.

### Data source

We used the Veterans Affairs (VA) Corporate Data Warehouse (CDW) to identify veterans with CTD. VA CDW is a national repository of longitudinal data of veterans who have received care at various VA medical centers since 1999.

### Case Definition

The CTD of interest included: RA, SLE, SSc, Sjogren’s, Dermatomyositis/Polymyositis (DM/PM), Overlap syndrome and other specified and unspecified CTD. The different ICD9 and ICD10 codes used for identification of CTD are included in **Appendix 1**. We included veterans who met the following criteria: (1) 18 years and older, (2) with at least two ICD9 or ICD10 codes for CTD separated by ≥6 months, (3) a rheumatology outpatient encounter within 6 months, and 4) diagnosis of CTD from Jan 1, 1999 to June 30^th^, 2022. A sample of 200 randomly selected patient charts were separately reviewed by two physicians to assess the sensitivity, specificity, positive predictive value, and negative predictive value of the CTD case definition.

To identify veterans who were diagnosed with ILD we used two categories. For Broad category we included all veterans with at least one ICD9 or ICD10 code for ILD (listed in **Appendix 2**) one year prior to diagnosis of CTD or anytime after CTD diagnosis. For Narrow category we included veterans who met the Broad Criteria and the following additional criteria within 6 months of ILD diagnosis: 1) pulmonary outpatient encounter, and 2) chest CT, and 3) pulmonary function testing. In order to ensure that the veterans had CTD related ILD, we excluded all patients who had ICD9 or ICD 10 diagnosis of ILD (**Appendix 3**) before the one year period preceding the diagnosis of CTD.

We selected ICD9 and ICD 10 codes for ILD from previously published studies with a few notable exceptions.^16–18^ The final codes we selected in Appendix 3 do not include ICD 9 codes 501 and 515 and ICD 10 code J61. We found during chart review that ICD codes 501 and J61, which are designated for pneumoconiosis due to asbestosis, were in many cases being used where the veteran had been exposed to asbestos but did not have pneumoconiosis. Similarly, ICD code 515 which is designated for post inflammatory fibrosis did not perform well during manual chart review similar to previously published study showing that almost 90% of patients with ICD code 515 did not have ILD.^19^ Therefore, ICD codes 501, 515 and J 61 were not used. We performed manual chart review of 65 randomly selected charts of veterans with at least one occurrence of ICD9 or 10 code for ILD in Appendix 3. The positive predictive value (PPV) of this approach for identifying ILD was 71%. From these ICD9 and ICD 10 codes, we selected codes that to identify patients with CTD-ILD in Appendix 2.

### Statistical Analysis

We calculated the prevalence of ILD (Broad and Narrow categories) within each CTD subtype in veteran patients and the timing of development of ILD after CTD diagnosis. Date of CTD diagnosis was the first occurrence of ICD9 or 10 code for CTD and date of ILD diagnosis was the first occurrence of ICD9 or 10 code for ILD. In those patients who developed ILD at or after diagnosis of CTD, we performed Kaplan Meier curves to determine the time to development of ILD after diagnosis of each of the different CTD subtypes. Cox regression analyses was performed to determine the risk of development of ILD adjusting for CTD subtype, age, gender at birth, race, and smoking status. Mortality was treated as a competing risk for cox regression analysis.

## Results

A total of 88,559 veterans with CTD were identified. The sensitivity, specificity, PPV, and negative predictive value of the case definition for CTD were 91%, 56%, 80% and 76% respectively on manual chart review. The average follow-up from CTD diagnosis was 8.8 (+/− 5.9) years. A majority of veterans with CTD were White (67.7%) followed by Blacks (17.5%) (**Table1**). The cohort was majority male (82.31%) with average age of 60.4 years (**Table 1**). RA was the most common CTD (n=72645; 82.03%), followed by SLE (n=6648; 7.51%) and Sjogren’s (n=3230; 3.65%) whereas DM/PM (n=1838; 2.08%) and Overlap syndromes (n=60; 0.07%) were the least prevalent diagnoses (**Table 1**).

**Table 1:** Baseline characteristics of veterans with connective tissue diseases.

| **Baseline Characteristics (N=88,559)** |  |
| --- | --- |
| **Age at diagnosis** | 60.4 (13.0) |
| **Male** | 72889 (82.31%) |
| **Female** | 15670 (17.69%) |
| **White** | 59,970 (67.7%) |
| **Black** | 15,534 (17.5%) |
| **Asian** | 622 (.7%) |
| **Rheumatoid Arthritis** | 72,210 (81.54%) |
| **Systemic Lupus Erythematosus** | 6,803 (7.68%) |
| **Systemic Sclerosis** | 2,110 (2.38%) |
| **Sjogren's** | 3,700 (4.18%) |
| **Overlap** | 114 (0.13%) |
| **Dermatomyositis/Polymyositis** | 1834 (2.07%) |
| **Other CTD** | 1788 (2.02%) |
| **ILD (Broad Definition)** | 8498 (9.6%) |
| **ILD (Narrow Definition)** | 3407 (3.85%) |

Using ILD Broad definition, a total of 8498 (9.59%) out of 88,559 veterans with CTD-ILD were identified. A total of 3407 (3.85%) out of 88,559 veterans met the Narrow definition of ILD diagnosis. Using the Broad definition, the prevalence of CTD-ILD was more common in SSc (42%) followed by DM/PM (21.7%), Overlap syndrome (19.3%), Other CTD (12.86%), SLE (11.22%), Sjogren’s (10.19%) and finally, RA (8.06%) (**Table 2**). Similar results were seen in the veterans who met the narrow criteria (results not shown). The mean age at CTD diagnosis for patients who developed ILD was lowest for patients with SLE at 49.71 (+/− 14.01) years and highest for patients with RA at 62.96 (+/− 10.21) years (**Table 3**). Among those who developed ILD anytime between 1 year prior to CTD diagnosis or after, the time to ILD diagnosis was lowest for patients with overlap syndrome at 2.15 (+-2.25) years and highest for RA at 6.01 (+/− 5.73) years (**Table 3**).

**Table 2:** Prevalence of Interstitial Lung Disease in various Connective Tissue Diseases.

| **CTD** | **Broad ILD** | **Narrow ILD** |
| --- | --- | --- |
| **RA**<br>**(N=72,210)** | 5,822<br>(8.06%) | 2,359<br>(3.26%) |
| **SLE**<br>**(N=6,803)** | 763<br>(11.22%) | 247<br>(3.63%) |
| **SSc**<br>**(N=2,110)** | 886<br>(41.99%) | 359<br>(17.01%) |
| **Sjogren's**<br>**(N=3,700)** | 377<br>(10.19%) | 152<br>(4.11%) |
| **Overlap**<br>**(N= 114)** | 22<br>(19.30%) | 7<br>(6.14%) |
| **DM/PM**<br>**(N=1834)** | 398<br>(21.70%) | 174<br>(9.49%) |
| **Other CTD**<br>**(N= 1788)** | 230<br>(12.86%) | 109<br>(6.1%) |

RA Rheumatoid Arthritis; SLE Systemic Lupus Erythematosus; SSc Systemic Sclerosis; DM/PM Dermatomyositis/Polymyositis
All percentages are row percentages.

**Table 3:** Mean age at diagnosis of Connective Tissue Diseases (CTD) and time to Interstitial Lung Disease (ILD) diagnosis in veterans with CTD-ILD using the Broad definition (N=8498).

| **CTD-ILD\*** | **Mean Age at CTD Diagnosis (SD)** | **Time to ILD Diagnosis in years (SD)** |
| --- | --- | --- |
| **RA (N=5,822)** | 62.96 (10.21) | 6.01 (5.73) |
| **SLE (N=763)** | 49.71 (14.01) | 5.18 (5.59) |
| **SSc (N=886)** | 57.08 (11.5) | 3.31 (4.30) |
| **Sjogren's (N=377)** | 59.56 (12.23) | 4.54 (5.30) |
| **Overlap (N= 22)** | 57.12 (11.58) | 2.15 (2.25) |
| **DM/PM (N=398)** | 56.35 (11.12) | 3.27 (4.83) |
| **Other CTD (N=230)** | 54.6 (13.70) | 3.64 (5.35) |

RA Rheumatoid Arthritis; SLE Systemic Lupus Erythematosus; SSc Systemic Sclerosis; DM/PM Dermatomyositis/Polymyositis
All percentages are row percentages.

Almost 29% of veterans meeting the Broad criteria developed ILD +/− 1 year of CTD diagnosis (**Table 4**). 632 (7.43%) of the Broad Criteria patients were diagnosed within a one-year period prior to the diagnosis of CTD (**Table 4**). The rest 7866 (92.56%) were diagnosed at or after CTD diagnosis (**Table 4**). 36% of patients with DM/PM, 33% with SSc, and 32% with Overlap developed ILD in the 1st year after CTD diagnosis compared to 20% of Sjogren’s and 18% of RA patients (**Table 4**). The details of race and gender for this cohort are detailed in **Tables E1 and E2**. Using the narrow criteria, 392 (11.5%) veterans were diagnosed with ILD within a one-year period prior to diagnosis of CTD and 3015 diagnosed at or after CTD diagnosis (data not published). A Kaplan Meier curve in the 7866 patients who developed ILD (using the Broad Definition) at or after CTD diagnosis shows a steady time to ILD diagnosis over the course of the follow up (**Figure 1**) and the curves are similar for all CTD categories except for Overlap syndrome (**Figure E1**). Similar Kaplan Meier curves were obtained using the narrow definition of ILD (**Figures E2 and E3**). The average time to diagnosis of ILD at or after diagnosis of CTD using the Broad definition was 5.84 (+− 5.58) years. The average time to ILD diagnosis at or after CTD diagnosis in the Narrow definition was 5.39 (+-5.4) years.

**Figure 1:**
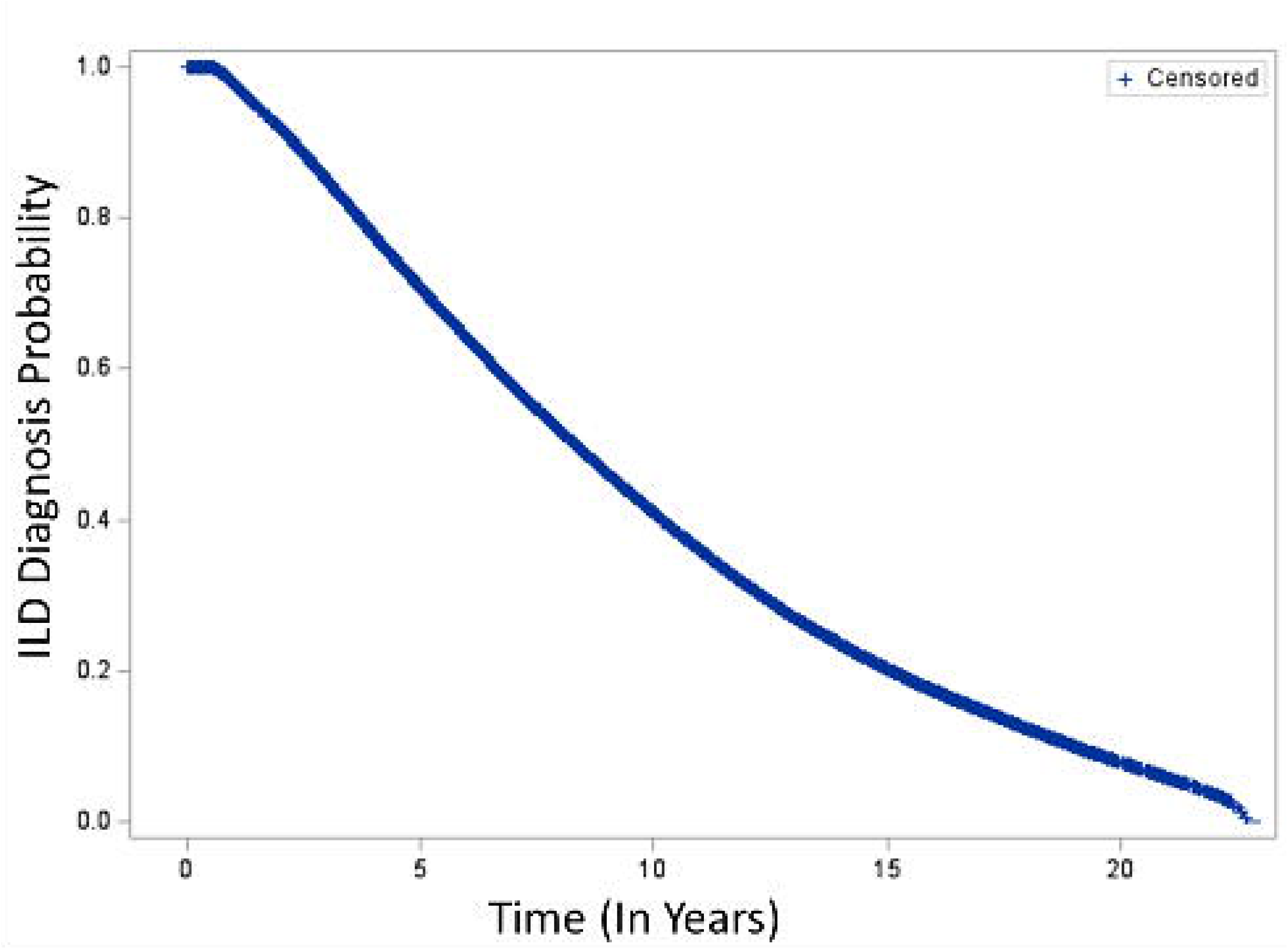
Kaplan Meier curve showing time to diagnosis of Interstitial Lung Disease (ILD) using the Broad definition in veterans at or after diagnosis of Connective Tissue Disease (CTD).

**Table 4:** Incidence of Interstitial Lung Diseases (ILD) using the Broad definition in Years from Connective Tissue Disease (CTD) Diagnosis.

| CTD | Years From CTD Diagnosis |  |  |  |  |  |  |  |  |  |  |  |
| --- | --- | --- | --- | --- | --- | --- | --- | --- | --- | --- | --- | --- |
|  | Pre CTD Dx* | 1<sup>st</sup> Yr | 2<sup>nd</sup> Yr | 3<sup>rd</sup> Yr | 4<sup>th</sup> Yr | 5<sup>th</sup> Yr | 6<sup>th</sup> Yr | 7<sup>th</sup> Yr | 8<sup>th</sup> Yr | 9<sup>th</sup> Yr | 10<sup>th</sup> Yr | >10 Yrs |
| **RA (5822)** | 345<br>(6) | 1063<br>(18) | 542<br>(9) | 437<br>(8) | 353<br>(6) | 336<br>(6) | 348<br>(6) | 287<br>(5) | 297<br>(5) | 231<br>(4) | 230<br>(4) | 1353<br>(23) |
| **SLE (763)** | 35<br>(5) | 201<br>(26) | 78<br>(10) | 61<br>(8) | 46<br>(6) | 37<br>(5) | 47<br>(6) | 33<br>(4) | 25<br>(3) | 22<br>(3) | 29<br>(4) | 149<br>(20) |
| **SSc (886)** | 85<br>(10) | 292<br>(33) | 110<br>(13) | 73<br>(8) | 64<br>(7) | 42<br>(5) | 36<br>(4) | 37<br>(4) | 29<br>(3) | 18<br>(2) | 19<br>(2) | 81<br>(9) |
| **Sjogren's (377)** | 55<br>(15) | 75<br>(20) | 36<br>(10) | 27<br>(7) | 32<br>(8) | 24<br>(6) | 14<br>(4) | 13<br>(3) | 22<br>(6) | 7<br>(2) | 12<br>(3) | 60<br>(16) |
| **Overlap (22)** | 3<br>(14) | 7<br>(32) | 0 | 3<br>(14) | 4<br>(18) | 2<br>(9) | 2<br>(9) | 1<br>(4) | 0 | 0 | 0 | 0 |
| **DM/PM (398)** | 58<br>(15) | 143<br>(36) | 39<br>(10) | 26<br>(7) | 17<br>(4) | 19<br>(5) | 16<br>(4) | 14<br>(3) | 8<br>(2) | 4<br>(1) | 6<br>(1) | 48<br>(12) |
| **Other CTD (230)** | 51<br>(22) | 57<br>(25) | 27<br>(12) | 12<br>(5) | 14<br>(6) | 6<br>(2) | 13<br>(6) | 7<br>(3) | 1<br>(0) | 4<br>(2) | 4<br>(2) | 34<br>(15) |
| **Total (8498)** | **632**<br>**(7)** | **1838**<br>**(22)** | **832**<br>**(10)** | **639**<br>**(8)** | **530**<br>**(6)** | **466**<br>**(5)** | **476**<br>**(6)** | **392**<br>**(5)** | **382**<br>**(4)** | **286**<br>**(3)** | **300**<br>**(4)** | **1725**<br>**(20)** |

\*Up to 1 year prior to diagnosis of CTD.
RA Rheumatoid Arthritis; SLE Systemic Lupus Erythematosus; SSc Systemic Sclerosis; DM/PM Dermatomyositis/Polymyositis
All values are numbers (percentages). All percentages are row percentages.

The adjusted Cox Regression analysis showed that the hazard of diagnosis of ILD in veterans with RA was 0.14 times compared to SSc (95%CI 0.13-0.15) adjusted for age, race, gender and smoking status (**Table 5**). The hazard for ILD for SLE, Sjogren’s and other CTD were similar at HR 0.2 when compared to SSc (**Table 5**). The hazard for ILD in overlap syndrome was not significant at 0.61 (95% CI 0.365-1.018). The hazard for ILD in DM/PM was 0.365 times that of SSc (95% CI 0.317 – 0.420) (**Table 5**). Of note, the smoking status had a large percentage of missingness and unknown categories (**Table E3**).

**Table 5:**
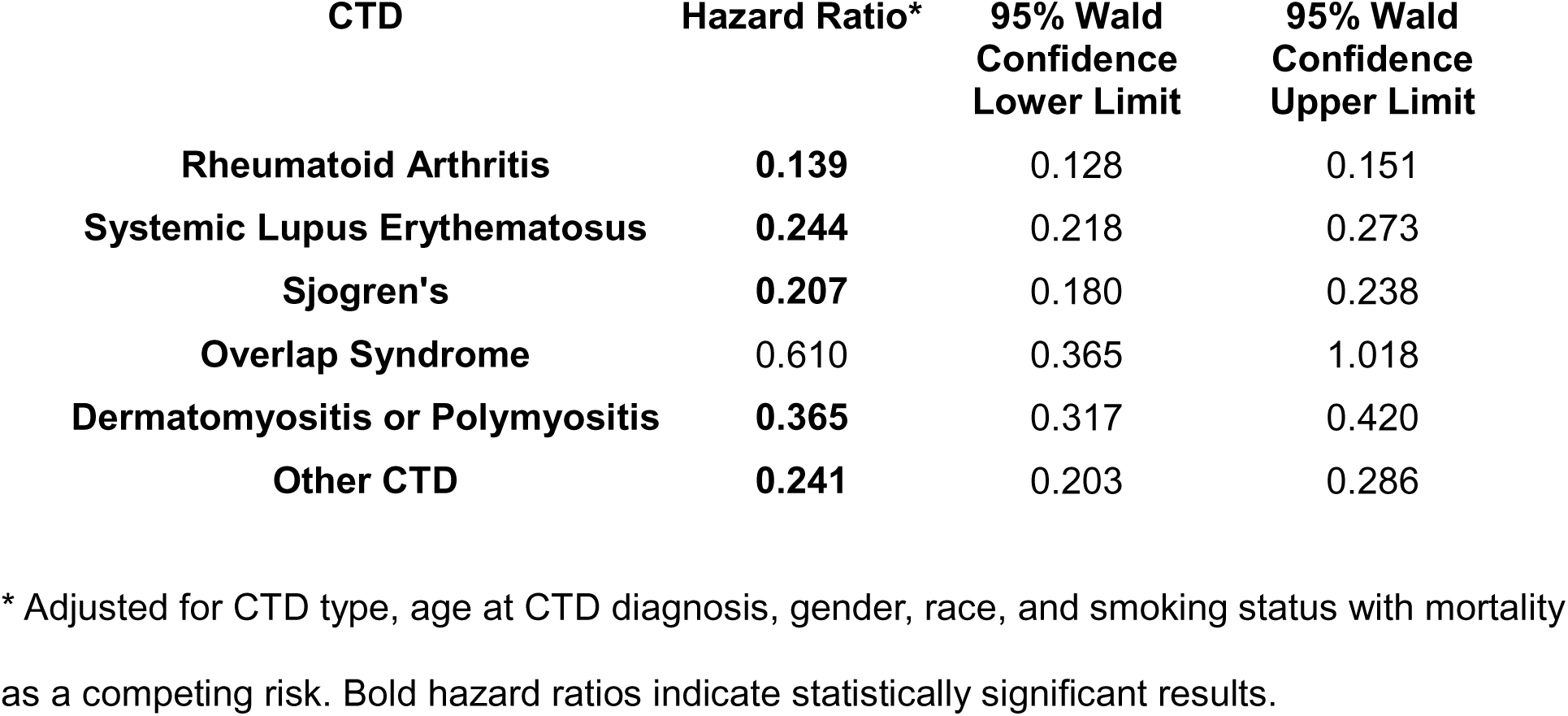
Cox Regression analysis of time to diagnosis of Interstitial Lung Disease (ILD) for various Connective Tissue Diseases (CTD) compared to Systemic Sclerosis in patients who developed ILD at or after CTD diagnosis using Broad definition (N=7866).

## Discussion

This study is the first comprehensive retrospective analysis of CTD and CTD-ILD in a national multicenter cohort of veterans. It demonstrates several notable clinical characteristics of veterans with CTD who were diagnosed with ILD either one year prior to CTD diagnosis or any time after. First, the prevalence of ILD in a national cohort of veterans with CTD was 9.6%. The prevalence was highest in SSc (42%) and lowest in RA at 8%. Second, 29% of patients developed ILD within +/− 1 year timeframe of CTD diagnosis. Third, the average time to diagnosis of ILD after a diagnosis of CTD is approximately 6 years. Collectively, these findings have important implications for the screening of ILD in veterans with CTD and for future research in CTD-ILD.

As previously noted, the prevalence of ILD varies depending on the CTD subtype, the study cohort, and the ascertainment method used in different studies to define ILD.^7^ Scleroderma had the highest prevalence of ILD of 42% in our cohort. This is similar to previous studies showing a prevalence of ILD in scleroderma ranging from 35-52%.^7,20,21^ Our cohort of veterans with CTD was predominantly composed of veterans with RA with the prevalence of ILD in veterans with RA of 8%. Prior studies have shown that prevalence of ILD in RA varies depending on the cohort and diagnostic modality used. In an old Australian study of patients with RA, 58% of patients had ILD with 76% of them having clinically silent disease.^22^ A more recent pooled analysis of 139 studies shows a prevalence of 11% in RA.^7^ Other studies show that ILD is detected on high resolution computed tomography (HRCT) in 60% of patients with RA but only 10% have clinically significant disease.^23^A study of US veterans with RA enrolled in a national registry found a 5.2% prevalence of physician coded ILD using limited ICD9 codes.^24^ Since our study used ICD 9 and 10 codes to identify ILD, we may only be identifying clinically significant disease in our cohort but we lack clinical data in our study to discriminate patients with symptomatic disease from those with subclinical disease. In patients with Rheumatoid Arthritis (RA), high titer anti-cyclic citrullinated peptide (anti-CCP), rheumatoid factor (RF), male sex, older age, smoking, and high disease activity confer higher risk for the development of ILD.^2,25^ Our cohort consisted of predominantly male patients with an average age of 60.4 years, both of which are known risk factors for the development of RA-ILD.^26,27^ Two European multidisciplinary studies investigating screening criteria for RA-ILD using a Delphi method proposed significantly different screening strategies: one suggesting a scoring system taking into account several risk factors while the other suggests waiting till decline in pulmonary function tests and development of symptoms.^28,29^ These recommendations were based on expert opinion and consensus given the lack of available evidence. The ACR guidelines suggest screening for patients with high titer anti-CCP or RF positivity who are deemed to be at high risk for ILD progression.^4^ The ERS/EULAR guidelines include additional criteria such as older age, smoking history, male sex, and high inflammatory markers and disease activity.^6^ However, both guidelines note the low level of evidence available to guide decision making in this field. Since a majority of RA patients may have findings of ILD on HRCT but otherwise asymptomatic and only 3-5% of patients have progressive disease, screening for early identification of ILD in all veterans with RA may not alter mortality.^4^

Early detection of ILD in patients with CTD should ideally help reduce mortality but it is also important to highlight patients with CTD-ILD experience a significant symptom burden that affects quality of life and causes financial strain. Symptoms not only impair daily function but also contribute to emotional distress, social isolation and loss of independence as noted in patient-centered studies.^30–32^ The economic impact is similarly significant. A systematic review of ILD-related economic data found that annual direct medical costs can range from $1,824 to $116,927 per patient and indirect costs, primarily from work productivity loss, added an additional $7,000–$11,000 annually per patient.^33^ In the United States, patients with rheumatoid arthritis–associated ILD (RA-ILD) incurred average annual healthcare costs ranging from $40,000 to $52,000.^34^ These findings highlight that CTD-ILD carries a significant toll and early detection of ILD could lead to earlier treatment and reduce symptom burden, progression and workplace productivity. Further research ideally in the form of prospective studies looking into cost effectiveness and more importantly mortality benefit in early screening of ILD in patients with CTD is needed.

The current guidelines conditionally recommend HRCT for initial screening at diagnosis for patients at risk for ILD followed by yearly rescreening per ACR (or more frequent if early in the disease course based on high risk autoantibody presence with PFTs).^4–6^ The ATS and ERS/EULAR guidelines do not make any recommendations about rescreening.^5,6^ While certainly screening with HRCT in the first year would capture at least 29% of ILD patients based on our study, using PFTs to rescreen patients annually or even more frequently may miss early disease since PFTs are not sufficiently accurate in identifying those with and without ILD.^6^ Given the low risk of radiation HRCT in the modern era, perhaps an annual CT scan in high risk patients may be effective screening strategy. Additionally, the current available guidelines do not make any recommendation on when to stop screening for ILD. As noted by our study, the incidence of ILD decreases below 10% after the first 5 years of CTD diagnosis (**Table 4**). Screening patients for up to 5 years after CTD diagnosis, which would capture ∼58% of ILD cases based on our results, is one possible strategy to limit screening and possible radiation exposure in patients.

The results of our study must also be interpreted within the context of limitations of the data source. First, the use of ICD 9 and 10 codes may have led to misclassification of veterans with CTD and ILD. While the manual chart review found that our CTD case definition had a PPV of 80% and our ILD algorithm had a PPV of 71%, we did not separately perform manual chart review of CTD-ILD using the broad or narrow definition. Additionally, the ICD10 code for overlap syndrome was not available until 2015, therefore, it accounts for only 0.7% of the ILD cases in our cohort which may be an underestimation. Second, the date of first occurrence of ICD9 or ICD 10 code of CTD or ILD may not be the date that these diagnoses were first identified or verified.

Therefore, the time to ILD diagnosis may be overestimated especially if the veteran was diagnosed with ILD outside of the VA. Third, many veterans are dual users of VA and non-VA health insurance. In these cases, our algorithm may fail to capture diagnosis and workup completed in the private medical sector. Thus, the true rate of CTD and ILDs among veterans reported in our study may be underestimated. However, a previous study has found that veterans with RA who utilize the VA tend to use the VA as the primary source of RA care.^35^ Finally, while the VA CDW captures longitudinal data for veterans’ care, antibody profile testing data is not easy to analyze. Over the course of the last 25 years, the VA’s various medical centers have used various lab test names for serum testing for RF, anti-CCP, anti-nuclear antibody, double stranded DNA, Sjogren’s antibodies, anti-topoisomerase antibodies, ribonucleoprotein antibody etc. Upon our initial analysis we found ∼2800 lab tests to identify the above antibodies. Additionally, these tests do not follow similar reporting for results making it difficult to accurately assess if the patients have a positive antibody result in the data. Inclusion of these antibody profiles would have strengthened the analysis of our data in selecting patients who would be at high risk for developing ILD but given the complexity of the data we were unable to include these markers in our study.

There is a critical need for large scale prospective studies to assess screening for ILD in patients with ILD and more importantly to assess the impact of screening on clinically relevant outcomes such as mortality which this current study was not designed to assess. Additionally, identifying a scoring system of patient factors within the high-risk categories who develop ILD and tend to develop progressive ILD could assist clinicians in identifying those patients who would benefit from early and more frequent screening for ILD.

## Data Availability

All data produced in the present study are available upon reasonable request to the authors

