## Appendix for "Interstitial Lung Diseases in a Longitudinal National Cohort of Veterans with Connective Tissue Diseases"

**Appendix 1: International Classification of Diseases (ICD) 9^th^ and 10^th^ Edition Codes for Connective Tissue Diseases**

| **Disease** | **ICD 9 and ICD 10 codes** |
| --- | --- |
| **Rheumatoid Arthritis (RA)** | 714.x, M05.1-M05.9, M06.0, M06.2, M06.3, M06.8, M06.9 |
| **Systemic Lupus Erythematosus (SLE)** | 710.0, M32 |
| **Scleroderma (SSc)** | 710.1, M34.x |
| **Sjogren’s** | 710.2, M35.0 |
| **Dermatomyositis or Polymyositis** | 710.3, 710.4, M33.x, M36.0 |
| **Other CTD** | 710.8, 710.9, M35.8, M35.9 |
| **Overlap Syndrome (ICD10 only introduced in 2015)** | M35.1 |

**Appendix 2: International Classification of Diseases (ICD) 9^th^ and 10^th^ Edition Codes for Interstitial Lung Diseases (ILD) Identification in Connective Tissue Diseases (CTD)**

**ICD 9 Codes**

516.30 Idiopathic Pulmonary fibrosis

516.32 Idiopathic non-specific interstitial pneumonia

516.33 Acute interstitial pneumonitis

516.34 Respiratory bronchiolitis interstitial lung disease

516.35 Idiopathic lymphoid interstitial pneumonia

516.36 cryptogenic organizing pneumonia

516.37 desquamative interstitial pneumonia

516.8 Other specified alveolar and parietoalveolar pneumopathies

516.9 Other nonspecific alveolar and parietoalveolar pneumopathies

517.1 Rheumatic Pneumonia

517.2 Lung Involvement in Systemic Sclerosis

*517.8 Lung involvement in other lung diseases classified elsewhere (these should have the following codes +/- 6 months)*

**ICD 10 Codes**

J84.1 Other interstitial pulmonary diseases with fibrosis

J84.2: LIP

J84.8 Other specified interstitial pulmonary disease

J84.9: Interstitial pneumonia NOS

M05.10-17: Rheumatoid lung disease with RA of specific site

M05.19: Rheumatoid lung disease with RA of multiple sites

M30.1 Polyarteritis with lung involvement (EGPA)

M32.13: Lung involvement in Lupus

M33.11: Other dermatopolymyositis with lung involvement

M33.21: Polymyositis with resp involvement

M33.91: Dermatopolymyosistis , unsp with resp involvement

M34.81: Systematic Sclerosis with lung involvement

M35.02: Sicca syndrome with lung involvement

**Appendix 3: International Classification of Diseases (ICD) 9^th^ and 10^th^ Edition Codes for Prior Diagnosis of Interstitial Lung Diseases (ILD)**

**ICD 9 Codes**

135 Sarcoidosis

495 Extrinsic allergic alveolitis

500 Coal workers' pneumoconiosis

502 Pneumoconiosis due to other silica or silicates

503 Pneumoconiosis due to other inorganic dust

504 Pneumonopathy due to inhalation of other dust

505 Pneumoconiosis, unspecified

508.1 Chronic and other pulmonary manifestations due to radiation

516.30 Idiopathic Pulmonary fibrosis

516.32 Idiopathic non-specific interstitial pneumonia

516.33 Acute interstitial pneumonitis

516.34 Respiratory bronchiolitis interstitial lung disease

516.35 Idiopathic lymphoid interstitial pneumonia

516.36 cryptogenic organizing pneumonia

516.37 desquamative interstitial pneumonia

516.4 Lymphanioleiomyomatosis

516.5 Adult pulmonary Langerhans cell histiocytosis

516.8 Other specified alveolar and parietoalveolar pneumopathies

516.9 Other nonspecific alveolar and parietoalveolar pneumopathies

517.1 Rheumatic Pneumonia

517.2 Lung Involvement in Systemic Sclerosis

*517.8 Lung involvement in other lung diseases classified elsewhere (these should have the following codes +/- 6 months)*

*+ 710.0 Lupus*

*+710.1 Systemic Sclerosis*

*+710.2 Sjogren’s*

*+710.3 Dermatomyositis*

*+710.4 Polymyositis*

*+714.81 Rheumatoid*

**ICD 10 Codes**

D86.0: Sarcoidosis of the lung

D86.2: Sarcoidosis of the lung and lymph nodes.

J60: Coal workers pneumoconiosis

J62: Pneumoconiosis due to Silicosis

J63: Pneumoconiosis due toother inorganic dusts

J64: Pneumoconiosis unspecified

J67: Hypersensitivity pneumonitis due to organic dusts

J70.1: Chronic and other pulmonary manifestations due to radiation

J70.3: Chronic drug-induced interstitial lung disorders

J70.4: Drug-induced interstitial lung disorders, unspecified

J82.81 Chronic Eosinophilic Pneumonia

J84.1 Other interstitial pulmonary diseases with fibrosis

J84.2: LIP

J84.8 Other specified interstitial pulmonary disease

J84.9: Interstitial pneumonia NOS

M05.10-17: Rheumatoid lung disease with RA of specific site

M05.19: Rheumatoid lung disease with RA of multiple sites

M30.1 Polyarteritis with lung involvement (EGPA)

M32.13: Lung involvement in Lupus

M33.11: Other dermatopolymyositis with lung involvement

M33.21: Polymyositis with resp involvement

M33.91: Dermatopolymyosistis , unsp with resp involvement

M34.81: Systematic Sclerosis with lung involvement

M35.02: Sicca syndrome with lung involvement

Table E1: Frequency table of race categories in veterans with Connective Tissue Disease Related Interstitial Lung Diseases (CTD-ILD) using the Broad definition (N=8498).

| **CTD-ILD** | **American Indian** | **Asian** | **Black or African American** | **Declined to answer** | **Native Hawaiian or Pacific Islander** | **Unknown** | **White** |
| --- | --- | --- | --- | --- | --- | --- | --- |
| **RA (N=5,822)** | 59 | 19 | 806 | 198 | 43 | 96 | 4334 |
|  | (1.06) | (0.34) | (14.51) | (3.56) | (0.77) | (1.73) | (78.02) |
| **SLE (N=763)** | 8 | 6 | 326 | 22 | 3 | 15 | 358 |
|  | (1.08) | (0.81) | (44.17) | (2.98) | (0.41) | (2.03) | (48.51) |
| **SSc (N=886)** | 23 | 8 | 241 | 38 | 2 | 14 | 498 |
|  | (2.79) | (0.97) | (29.25) | (4.61) | (0.24) | (1.7) | (60.44) |
| **Sjogren’s (N=377)** | 6 | 3 | 107 | 12 | 4 | 5 | 232 |
|  | (1.63) | (0.81) | (29) | (3.25) | (1.08) | (1.36) | (62.87) |
| **Overlap (N= 22)** | 0 | 0 | 10 | 0 | 0 | 0 | 12 |
|  |  |  | (45.45) |  |  |  | (54.55) |
| **DM/PM (N=398)** | 2 | 4 | 165 | 10 | 5 | 9 | 184 |
|  | (0.53) | (1.06) | (43.54) | (2.64) | (1.32) | (2.37) | (48.55) |
| **Other CTD (N=230)** | 6 | 2 | 91 | 7 | 2 | 3 | 113 |
|  | (2.68) | (0.89) | (40.63) | (3.13) | (0.89) | (1.34) | (50.45) |
| **Total** | 104 | 42 | 1746 | 287 | 59 | 142 | 5731 |
|  | (1.28) | (0.52) | (21.53) | (3.54) | (0.73) | (1.75) | (70.66) |
| **Frequency Missing = 387** | | | | | | | |

RA Rheumatoid Arthritis; SLE Systemic Lupus Erythematosus; SSc Systemic Sclerosis; Sjogren’s; DM/PM Dermatomyositis/Polymyositis

Values are number (percentages). All percentages are row percentages.

Table E2: Frequency table for gender categories in veterans with Connective Tissue Disease Related Interstitial Lung Diseases (CTD-ILD) using Broad definition (N=8498).

| **CTD-ILD*** | **Male** | **Female** |
| --- | --- | --- |
| **RA (N=5,822)** | 5,385 (92.49%) | 437 (7.51%) |
| **SLE (N=763)** | 428 (56.91%) | 335 (43.91%) |
| **SSc (N=886)** | 698 (78.78%) | 188 (21.22%) |
| **Sjogren’s (N=377)** | 233 (61.80%) | 144 (38.20%) |
| **Overlap (N= 22)** | 17 (77.27%) | 5 (22.73%) |
| **DM/PM (N=398)** | 30 (85.43%) | 58 (14.57%) |
| **Other CTD (N=230)** | 172 (74.78%) | 58 (25.22%) |

RA Rheumatoid Arthritis; SLE Systemic Lupus Erythematosus; SSc Systemic Sclerosis; Sjogren’s; DM/PM Dermatomyositis/Polymyositis

All percentages are row percentages.

Table E3: Smoking status in veterans with Connective Tissue Disease Related Interstitial Lung Diseases (CTD-ILD) using the broad category (N=7866).

| **Smoking Status** | **Number** |
| --- | --- |
| **Current Smoker** | 1961 (27.62%) |
| **Former Smoker** | 1817 (27.59 %) |
| **Never Smoker** | 1672 (23.55%) |
| **Unknown** | 1650 (23.24%) |
| **Missing = 766** | |

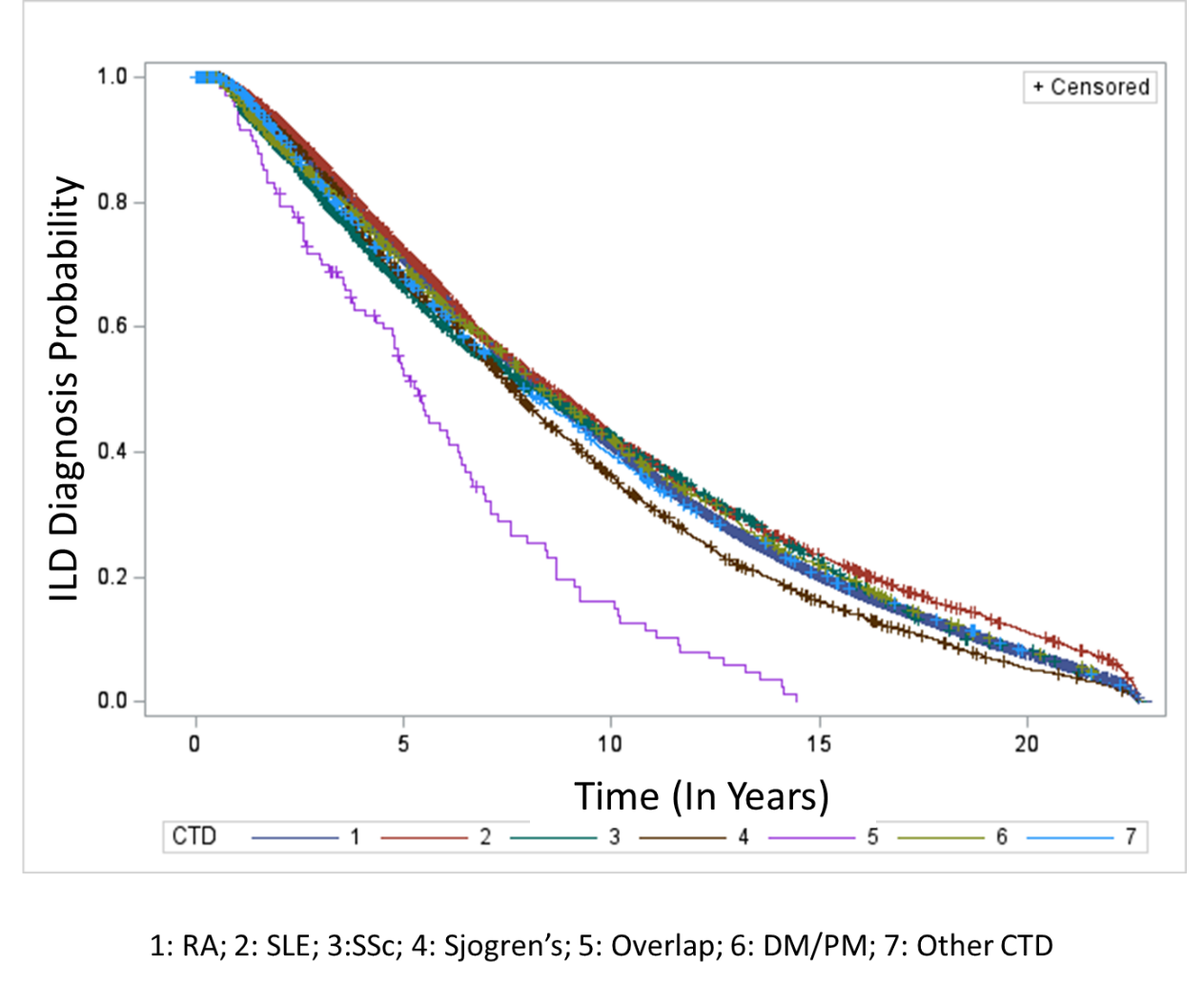

Figure E1: Kaplan Meier curve showing time to diagnosis of Interstitial Lung Disease (ILD) using the Broad definition in veterans at or after diagnosis of Connective Tissue Disease (CTD) for different CTD types. RA Rheumatoid Arthritis; SLE Systemic Lupus Erythematosus; SSc Systemic Sclerosis; Sjogren’s; DM/PM Dermatomyositis/Polymyositis

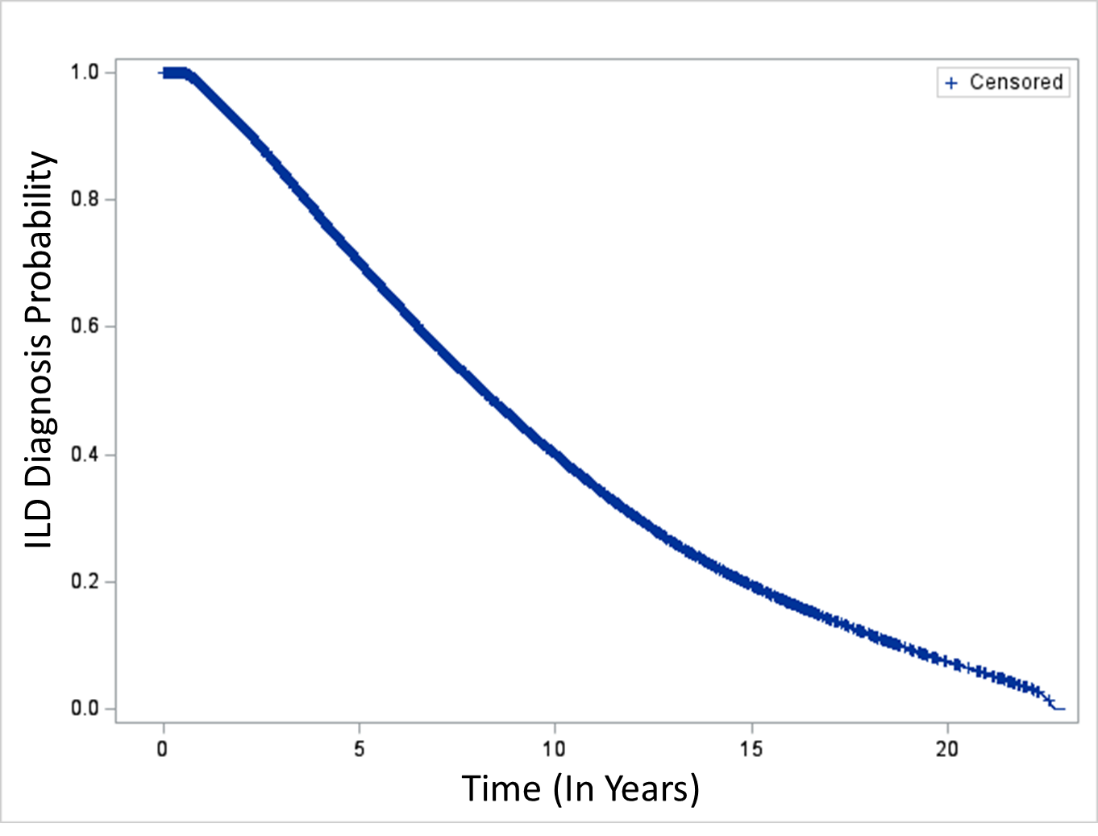

Figure E2: Kaplan Meier curve showing time to diagnosis of Interstitial Lung Disease (ILD) using the Narrow definition in veterans at or after diagnosis of Connective Tissue Disease (CTD).

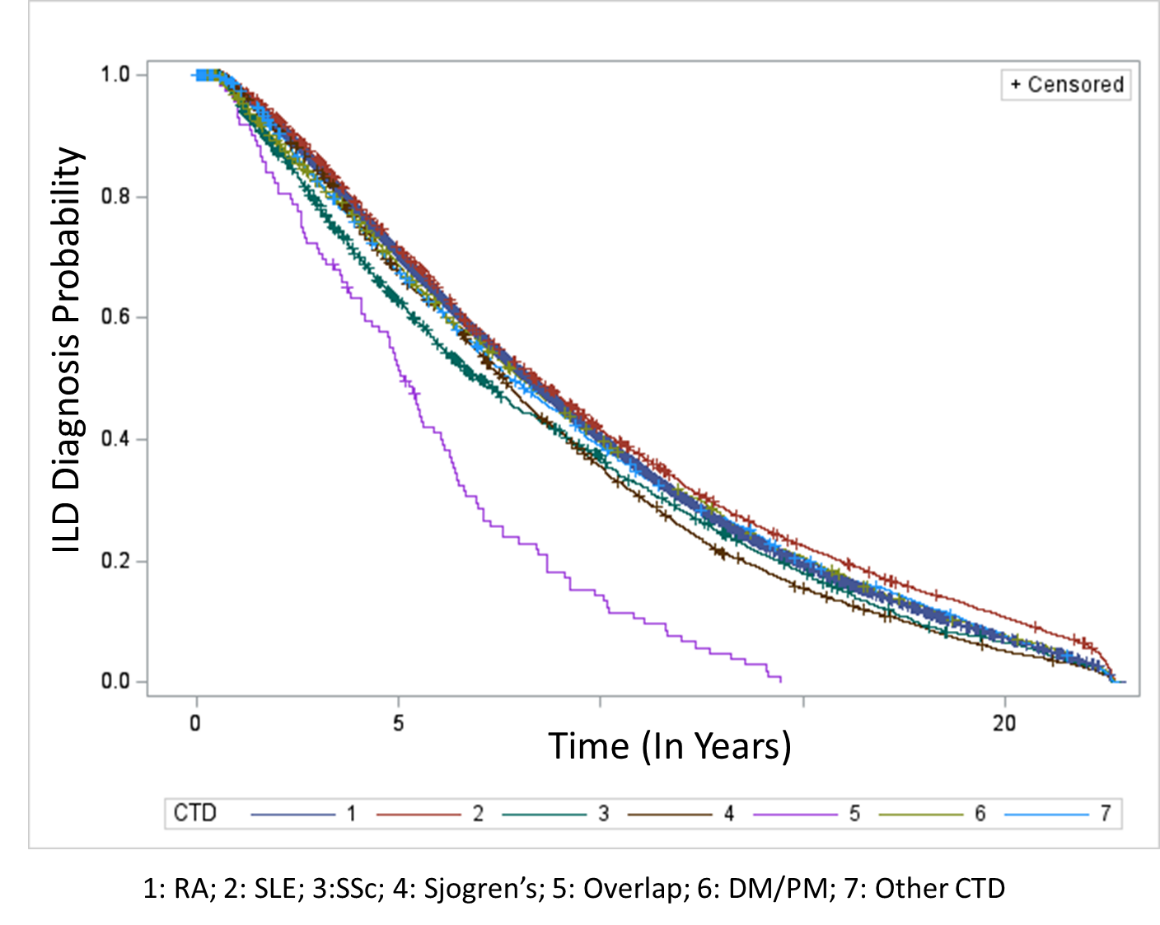

Figure E3: Kaplan Meier curve showing time to diagnosis of Interstitial Lung Disease (ILD) using the Narrow definition in veterans at or after diagnosis of Connective Tissue Disease (CTD) for different CTD types. RA Rheumatoid Arthritis; SLE Systemic Lupus Erythematosus; SSc Systemic Sclerosis; Sjogren’s; DM/PM Dermatomyositis/Polymyositis
